# Long-read sequencing resolves a complex structural variant in *PRKN* Parkinson’s disease

**DOI:** 10.1101/2023.08.14.23293948

**Authors:** Kensuke Daida, Manabu Funayama, Kimberley J Billingsley, Laksh Malik, Abigail Miano-Burkhardt, Hampton L. Leonard, Mary B. Makarious, Hirotaka Iwaki, Jinhui Ding, J. Raphael Gibbs, Mayu Ishiguro, Hiroyo Yoshino, Kotaro Ogaki, Genko Oyama, Kenya Nishioka, Risa Nonaka, Wado Akamatsu, Cornelis Blauwendraat, Nobutaka Hattori

## Abstract

**Background:** *PRKN* mutations are the most common cause of young onset and autosomal recessive Parkinson’s disease (PD). *PRKN* is located in FRA6E which is one of the common fragile sites in the human genome, making this region prone to structural variants. However, complex structural variants such as inversions of *PRKN* are seldom reported, suggesting that there are potentially unrevealed complex pathogenic *PRKN* structural variants.

**Objectives:** To identify complex structural variants in *PRKN* using long-read sequencing.

**Methods:** We investigated the genetic cause of monozygotic twins presenting with a young onset dystonia-parkinsonism using targeted sequencing, whole exome sequencing, multiple ligation probe amplification, and long-read. We assessed the presence and frequency of complex inversions overlapping *PRKN* using whole-genome sequencing data of AMP-PD and UK-Biobank datasets.

**Results:** Multiple ligation probe amplification identified a heterozygous exon 3 deletion in *PRKN* and long-read sequencing identified a large novel inversion spanning over 7Mb, including a large part of the coding DNA sequence of *PRKN*. We could diagnose the affected subjects as compound heterozygous carriers of *PRKN*. We analyzed whole genome sequencing data of 43,538 participants of the UK-Biobank and 4,941 participants of the AMP-PD datasets. Nine inversions in the UK-Biobank and two in AMP PD were identified and were considered potentially damaging and likely to affect *PRKN* isoforms.

**Conclusions:** This is the first report describing a large 7Mb inversion involving breakpoints outside of *PRKN*. This study highlights the importance of using long-read whole genome sequencing for structural variant analysis in unresolved young-onset PD cases.

## Introduction

Parkinson’s disease (PD) is the second most common neurodegenerative disease next to Alzheimer’s disease, developing with symptoms such as bradykinesia, resting tremor, rigidity and postural instability.^1^ PD is typically divided into two categories: young-onset PD and late-onset PD. The cut-off age that defines young onset PD has been ambiguous, but it has recently been defined by the Movement Disorder Task Force as any age at onset before the age of 50.^2^ Genetics plays a vital role in young onset PD and the *parkin RBR E3 ubiquitin-protein ligase* (*PRKN)* gene is the most frequent causative gene in young onset PD and autosomal recessive PD,^3^ which were initially identified in a Japanese family.^4^ *PRKN* encodes the parkin protein which is an E3 ubiquitin-protein ligase that maintains mitochondrial function by removing dysfunctional mitochondria through autophagy. This process heavily involves *PINK1,* where *PINK1* mutations are the second most common cause of young-onset PD and autosomal recessive PD.^5^

*PRKN* is located on chromosome 6q25.2-27, containing 12 coding exons and spanning a total of 1.3Mb. The region of *PRKN* is within FRA6E, which is the third most frequently observed common fragile site of the human genome.^6,7^ Common fragile sites are specific loci that preferentially exhibit gaps and breaks on metaphase chromosomes following partial inhibition of DNA synthesis.^8^ The central core of FRA6E is located in exon 3 to 8 of *PRKN*, which is the known mutation hot spot of *PRKN.*^7^ To date, over a hundred pathogenic *PRKN* mutations including point mutation and structural variants (SVs) are reported in the Movement Disorder Society Genetic mutation database (https://www.mdsgene.org/). Biallelic *PRKN* mutation accounts for an estimated 4.3 % and 8-15 % of sporadic and familial young onset PD.^9,10^ The role of heterozygous *PRKN* variants in PD is controversial, where some reports show increased risk of carrying a single damaging variant and others report no effect.^11–16^ It is worth noting that the reported associations between heterozygous *PRKN* and PD in those studies may be influenced by a potential second unrevealed and complex to identify mutation that affects the function of *PRKN*.

Biallelic *PRKN* patients typically present young or juvenile-onset Parkinsonism with levodopa responsiveness, dystonia, dyskinesia, and motor fluctuations, but without cognitive decline, autonomic symptoms, and psychotic symptoms.^17^ The pathology of the brain with *PRKN* variants is characterized by neuronal loss in substantia nigra pars compacta often without Lewy body.^18^

Long-read sequencing has been an emerging sequencing technique where reads longer than 10,000 bp can reliably be sequenced, which is >30 times larger than conventional short-read sequencing.^19^ Not only do longer reads (10kb+) improve *de novo* assembly and mapping, but they are better able to detect SVs because they can span repetitive or other complex regions of the genome. For identifying SVs in PD specifically, a recent study that compared matched long and short sequencing read data from PD cases highlighted that most SVs in the human genome are likely undetectable with short-read data alone (∼84%).^20^ Long-read sequencing is a powerful tool for identifying potential causal variants that were previously undetectable using other sequencing technologies.

Here we describe a *PRKN* family with two affected monozygotic twins presented clinically with dystonia Parkinsonism. No other family members had been diagnosed with PD or any other neurological diseases. Initial genetic testing identified a single *PRKN* mutation despite showing a clear typical *PRKN* clinical presentation. After extensive follow-up using long-read sequencing we identified an additional complex SV at the *PRKN* locus explaining the *PRKN*-PD phenotype.

## Methods

### Study participants

The study was approved by the ethics committee of Juntendo University, Tokyo, Japan, and all participants provided written informed consent to participate in the genetic research described in this study. PD was clinically diagnosed according to standard clinical criteria.^21,22^ DNA was extracted from peripheral blood by the standard protocol using QIAamp DNA Blood Maxi Kit (QIAGEN, Venlo, Netherlands).

### Targeted panel sequencing using short read whole exome sequencing

Targeted panel sequencing and whole exome sequencing (WES) were performed in II-3, III-1 and III-2 (Fig. 1A). The method of targeted panel sequencing for PD-related genes has been previously reported.^23^ Libraries for WES were prepared with SureSelect Human All Exon V6 kit (Agilent Technologies Santa Clara, CA, USA). Libraries were sequenced using the Illumina Hiseq1500 (Illumina, San Diego, CA, USA). Picard was used to mark duplicates, and variants were called according to GATK v.4.1.3.0 pipeline.^24^ Annotation was conducted by Annovar.^25^ Variants were filtered according to the following criteria: base quality score, location in exons or splice sites, and the allele frequency (gnomAD) smaller than 0.001.^26^

**Figure 1.**
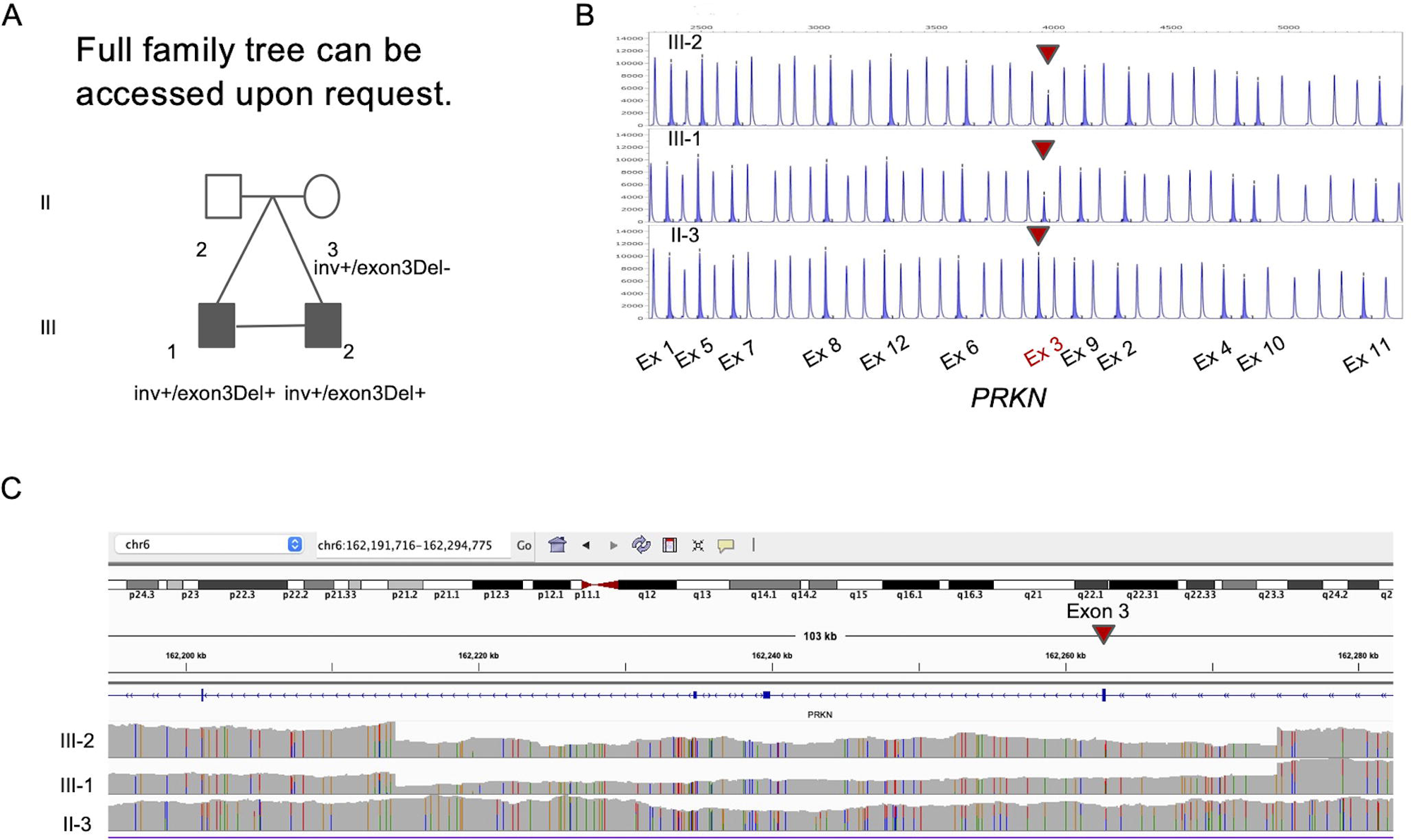
Family Pedigree and Genetic test results of the twins family. (A) Family pedigree with *PRKN* inv/exon 3 deletion. Squares, men; circles, women; oblique lines, deceased; black squares, clinically diagnosed with PD (B) MLPA result for *PRKN* exons (C) Screenshot from IGV presenting with Exon 3 deletion in *PRKN*.

### Multiplex Ligation-dependent Probe Amplification

Copy number variants (CNV)s in *PRKN* and *SNCA* were analyzed using multiplex Multiplex Ligation-dependent Probe Amplification (MLPA) with SALSA MLPA Probemix P051/P052 Parkinson probe mix (MRC-Holland, Amsterdam, the Netherlands). MLPA experiments were done according to the manufacturer’s instructions.

### Oxford Nanopore Technologies long-read sequencing

We used the DNA prepared for the short-read sequencing as starting material for the long-read sequencing. Sequencing was prepared according to our protocol reported previously.^27^ In brief, DNA samples were sized using the Femto Pulse (Agilent Technologies Santa Clara, CA, USA). DNA underwent through a size selection step using the Circulomics Short Read Eliminator Kit (SS-100-101-01) to remove fragments up to 25kb. Libraries were prepared using the Kit V14 Ligation sequencing kit from ONT and sequenced using PromethION for 72 hours on a R10.4.1 flow cell (Oxford Nanopore Technologies, Oxford, UK). Base calling was performed by Guppy v6.3.8^28^ and Winnowmap v2.0.3^29^ was used to map the reads to the GRCh38 reference genome. Sniffles v2.0.7^30^ was used for calling SVs (using –tandem-repeats option). SVs were annotated by AnnotSV v3.1.1.^31^

### Inversion breakpoint region sequence analysis

To confirm the breakpoints of the inversion that were identified by long-read sequencing, we amplified the breakpoint regions using PCR by primers specifically designed by Primer 3 to detect the inversion (Supplementary Table 1). To consider how the breakpoints occur, we performed breakpoint region sequence analysis as described previously.^32,33^ We obtained the sequence of 100 bp upstream and downstream of the breakpoints from the UCSC genome browser. (https://genome.ucsc.edu/index.html). Repeat information was obtained by RepeatMasker (Smit, AFA, Hubley, R & Green, P. RepeatMasker Open-4.0. 2013-2015, http://www.repeatmasker.org). Sequence similarity around the breakpoints was checked using EMBOSS Needle^34^. We also used palindrome software to identify sequences with potential to cause stem loops (https://emboss.bioinformatics.nl/cgi-bin/emboss/palindrome).

### Illumina short read whole genome sequencing

To replicate the findings in long-read sequencing, we conducted short-read whole genome sequencing (WGS) to III-2. WGS was performed by library preparation using the VAHTS Universal Pro DNA Library Prep Kit from Illumina and sequencing was performed using a Illumina Novaseq 6000 sequencer at 150 bp paired-end sequencing. The estimated data generated per sample was >90 Gb resulting in >30 coverage. Reads were aligned by Burrows-Wheeler Alignment tool v0.7.17.^35^ Short-read WGS was analyzed in the same framework as the short-read WES which is written above. SVs were called using Manta v1.6.0.^36^

### Replication using Short read Whole genome sequencing datasets

As the inversion was also identified after targeted re-analysis of the short-read sequencing, we used the UK Biobank short-read WGS data, AMP-PD cohort to examine the frequency of the inversions of *PRKN* in controls and cases. AMP-PD cohort is a dataset of short-read WGS which are described previously.^16^ We called the SVs using Manta^36^ and used the output of diploidSV.vcf.gz. We used the Manta-called scored SV and indel candidates In UK Biobank (Field ID 23350). Using both datasets, the variants that matched the following criteria remained for further analysis: 1) variant type is inversion, 2) the inversion affects the transcript of *PRKN*, and 3) the size of the SV is within 50 bp to 10 Mb. The filtered variants were confirmed visually using IGV to filter false positive inversions (called as inversions for several chromosomes).^37^

### Data Sharing

UK Biobank data is available upon application at the UK Biobank website (https://www.ukbiobank.ac.uk/). AMP-PD data is available upon application at the AMP-PD website (https://amp-pd.org/).

## Results

Here we report monozygotic twins presented with a young onset dystonia Parkinsonism phenotype (Fig. 1A). They were born without any problems during the pregnancy. Their family is of Japanese ancestry and has no reported consanguinity (Fig. 1A). The clinical symptoms of the affected twins at their 30’s are summarized in Supplementary Table 2. Details of the clinical symptoms, full family tree, and Supplementary Table 2 is available upon request to the corresponding authors.

### Clinical features of the older brother (III-1)

The age at onset was at 10’s, the initial symptom was spastic gait. He gradually developed bradykinesia and postural instability. Levodopa response was positive and aiding with symptoms, and the amount of levodopa increased to 500 mg at late 20’s. Wearing off had gradually made his daily life difficult. At mid 30’s he started to take rotigotine. Rotigotine improved his symptoms; however he started to show impulse control disorder, mainly gambling. As the wearing-off worsened, he underwent a subthalamic nucleus deep brain stimulation operation at late 30’s. He did not present cognitive decline throughout the disease’s progress. 123-Iodine Metaiodobenzylguanidine myocardial scintigraphy showed normal heart-to-mediastinum ratio (early 2.39, delay 2.99). Brain magnetic resonance imaging (MRI) was normal, and DAT-SPECT showed decreased uptake in both basal ganglia (Fig. 2A). Brain SPECT with N-isopropyl-p[123I]-iodoamphetamine (IMP-SPECT) revealed reduced basal ganglia.

**Figure 2.**
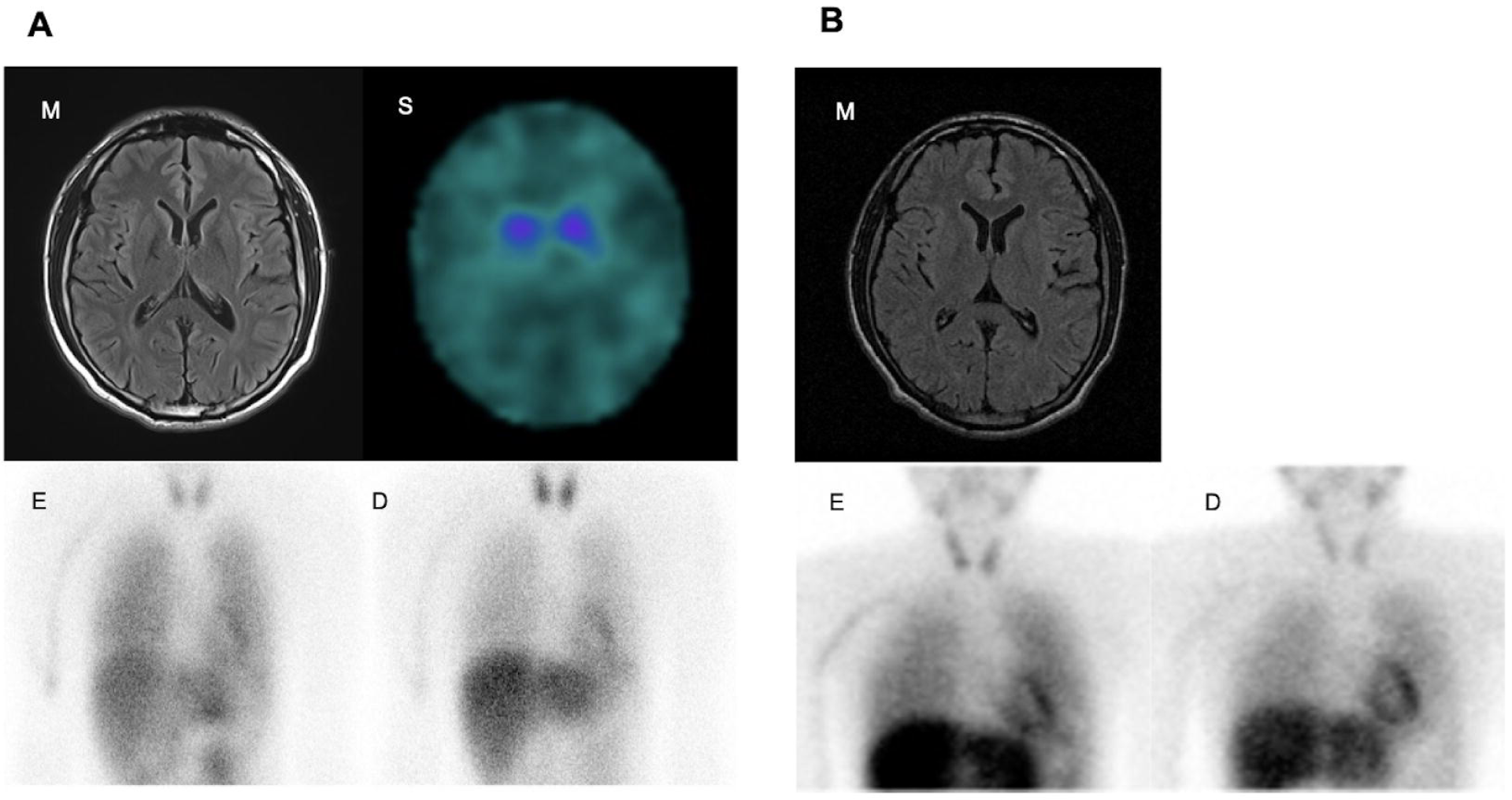
Clinical Images. (A) Images for III-1. (B) Images for III-2. (m) Brain MRI (s) DAT-SPECT. MIBG myocardial scintigraphy on early phase (e) and delayed phase (d).

### Clinical features of the younger brother (III-2)

The age at onset was same as the younger brother. He presented with walking difficulties and right lower limb dystonia. At the first visit to the neurological clinic at early 20’s, he was considered spastic paraplegia. Levodopa showed significant effects on dystonia. The amount of levodopa increased gradually; he took 500mg of levodopa at late 30’s. A few years later, he presented with diurnal variation of dystonia, bradykinesia, dyskinesia, and posture instability. The Hoen and Yahr scale was zero at the on phase, three at the off phase. He had started to take levodopa 50 mg frequently (in total, max 500 mg/day), which helped to control his symptoms. Since then, he has been in reasonable control, presenting almost no symptoms at the latest visit at early 40’s. Metaiodobenzylguanidine myocardial scintigraphy showed normal heart-to-mediastinum ratio (early = 2.68, delay= 2.98). Brain MRI was normal, and IMP-SPECT did not show aberrant flow distribution (Fig. 2B).

### Exploring the genome for potential causal variants

DNA was available for the affected twins (III-1, III-2) and mother (II-3) but was not available for the father (II-2). Targeted panel sequencing against known PD-related genes and WES identified a heterozygous *PRKN* variant c.814C>A (p.L272I, rs141366047) in the affected subjects. The allele frequency of c.814C>A is 0.014 by 38KJPN in jMorp,^38^ one of the largest genomic databases including the Japanese population, suggesting it is a relatively common SNP in the Japanese population and therefore is not likely to be pathogenic. MLPA revealed a heterozygous deletion spanning Exon 3 of *PRKN* in the affected twins but not in the mother (Fig. 1B). No other variants were identified to be of interest.

Since the affected twins presented typical young onset PD *PRKN* phenotype, we suspected that there might be a complex undetected variant in *PRKN*, which could be missed by short-read WES and MLPA. Therefore we generated long-read sequencing data for the twins and the mother using Oxford Nanopore Technologies Long-read sequencing. Sample DNA QC results were good enough to perform long-read sequencing (Supplementary Figure 4 and Supplementary Table 4). The overall data output for the samples ranged from 120-130 Gb (∼38X coverage assuming 3.1Gb genome) and the average read N50s were 19kb (Supplementary Table 3). Long-read sequencing confirmed the 60,138 bp deletion (hg38) including exon 3 of *PRKN* (c.(171+1_172-1)_(412+1_413-1)del) in the affected twins but not in the mother, which is concordant with the result of MLPA (Fig. 1C). Additionally long-read sequencing revealed a second mutation, a heterozygotic inversion spanning approximately 7,425,905 bp (hg38), involving exon 1 to 11 in affected twins and mother (NC_00006.12:g.161351957_168777862inv). The proximal breakpoint junction was in the intron 11 of *PRKN*, which is predicted to remove exon 12 from the transcript and therefore resulting in a non-functioning transcript. Importantly as expected based on MLPA there was no change in the peak of exon 12 in all samples (Fig. 1B). Each of the breakpoints did not have insertion sequences. The allele frequency of this inversion is unknown since no inversion including *PRKN* has been reported in JSV1 from jMorp, a long-read WGS database in the Japanese population ^38^. Other than the two *PRKN* variants reported above, we did not identify other potential SVs in PD-related genes. PCR confirmed both breakpoints of the inversion (Supplementary Figure. 1) and additional short-read WGS confirmed the deletion and inversion (Supplementary Figure. 2 and 3).

### Breakpoint region sequence analysis of the PRKN inversion

To consider how the breakpoints occur for this inversion, we performed breakpoint region sequence analysis. Based on the inversion (Fig 3A), there is a short interspersed nuclear element (SINE) transposable element (148 bp in length) 96 bp upstream of the 5’ breakpoint and a long interspersed nuclear element transposable element (245 bp in length) 178 bp downstream of the 3’ break point. Sequence similarity was 37.1% in 100 bp up and downstream of the both break points. There was no palindromic sequence around both the breakpoints. Both breakpoints for the deletion of exon 3 (chr6:162,214,329-162,274,466del) were blunt ends. 3’ breakpoint is in the SINE sequence. Sequence similarity was 42.3% in 100 bp up and downstream of the both break points.

**Figure 3.**
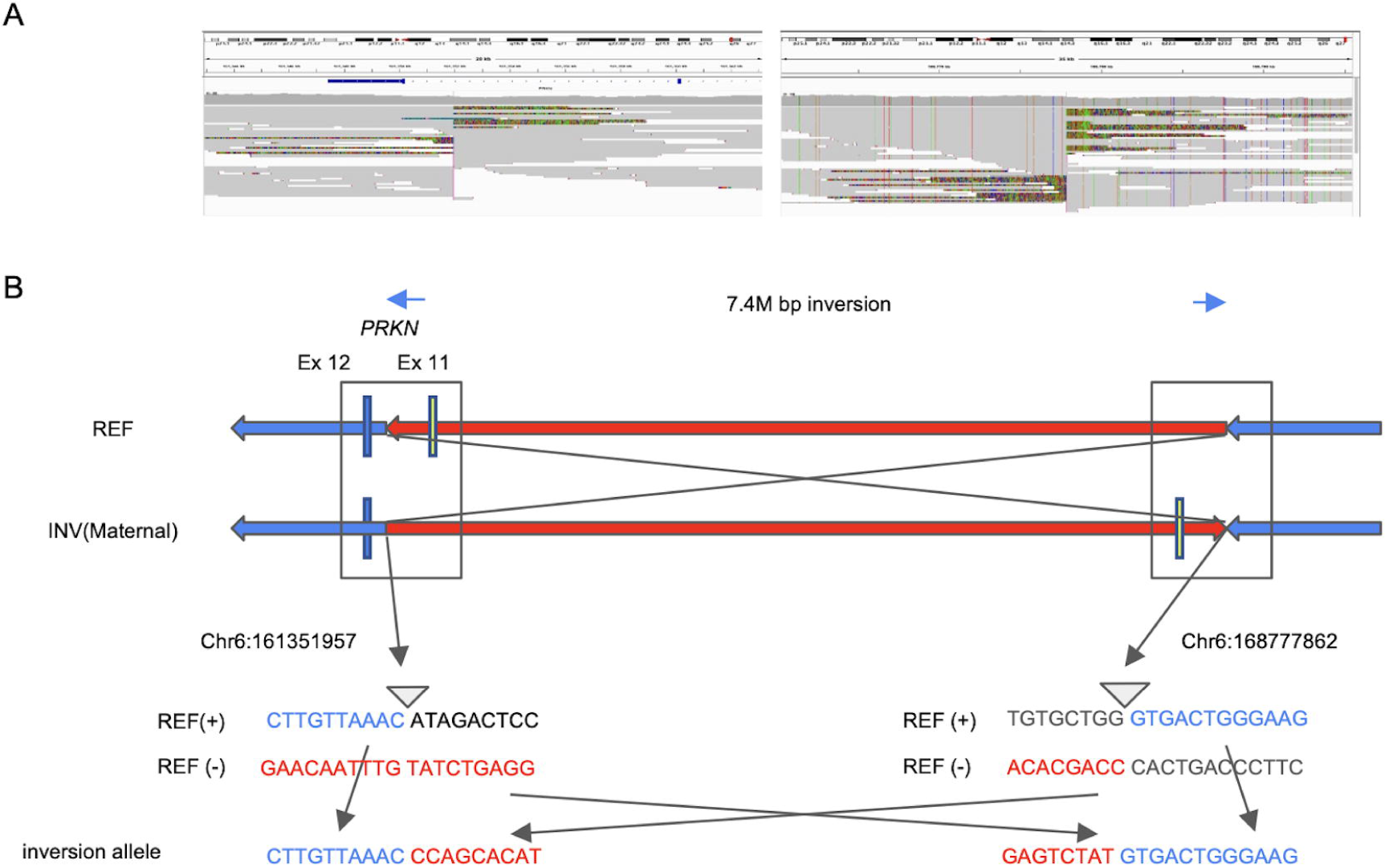
Detailed description of the identified inversion. (A) Screenshot from IGV presenting breakpoints of 7M bp inversion including *PRKN.* The left figure represents the 5’ break point. The breakpoint is located in intron 11 of *PRKN*. The right figure shows the 3’ breakpoint. (B) Schematic representation of the inversion. The upper panel shows the overall picture of the inversion. The lower panel shows the sequence around both breakpoints. 5’ breakpoint Chr6:161351957, 3’ breakpoint Chr6:168777862. REF = reference; INV = inversion

### Assessing the frequency of large inversions involving *PRKN* using short read WGS from AMP-PD and UK Biobank

Next we wanted to assess the frequency of large inversions in the *PRKN* genomic region. We used Manta which is a short read structural variant caller on two large datasets for this: the AMP-PD dataset which includes 3403 samples (1131 controls and 2272 PD cases) after QC and a subset of the UK Biobank cohort which has WGS data available on the online RAP platform including 43,858 participants and including 259 PD cases (based on ICD10 code: G20, Parkinson’s disease). In the AMP-PD dataset, we identified 11 inversions in 17 subjects and two of these 11 inversions included a coding exon of PRKN (NM_004562) the others were intronic (Supplementary Table 5). These two inversions affecting exons were called from the same PD subject whose age at diagnosis was also early onset PD with reported age of onset in 40’s. After about 20 years since his onset, he took only 280mg levodopa per day and his MDS-UPDRS scores were not high (MDS-UPDRS I/II/III/IV; 7/1/21/0), showing slow progression of his disease. His Montreal cognitive Score Assessment score was 29 points suggesting no cognitive decline. Those clinical features were compatible with *PRKN*-PD. However because this data is based on short-read sequencing we cannot distinguish whether these two inversions are on the same allele or not. We also confirmed he does not have any susceptible pathogenic point mutation in *PRKN* from short-read data.

In the UK Biobank cohort, 43,858 subjects were included and 31 inversions in 46 subjects were found. Nine of the identified inversions included one or more *PRKN* exons and the other 22 were intronic and did not affect a coding exon. Using IGV, one of the nine exon-including inversions was considered to be false positive since mate reads were mapped to several chromosomes (Supplementary Figure 5). All the inversions including exon in UK Biobank were identified from non PD subjects. Two inversions in AMP-PD and nine inversions in the UK Biobank were considered potential inversions to affect *PRKN* transcripts.

## Discussion

In this study, we identified two compound heterozygous SVs in *PRKN* in monozygotic twins with PD. The age of onset of the twins was in their teens and the clinical symptoms of the twins were very similar and typical for young onset PD, suggesting a genotype-phenotype correlation. However only a heterozygotic deletion of exon 3 of *PRKN* was initially identified by short-read sequencing WES and MLPA. Long-read sequencing revealed a large inversion expanding 7.4M bp with 5’ breakpoint in intron 11 of *PRKN*, removing exon 12 from *PRKN* transcripts. In agreement with recent studies,^20^ our study highlights that long-read sequencing can be used to identify new SVs in the PD population, especially in *PRKN*-PD.

The age of onset of the affected twins was in their teens, which was younger compared to the median age (31 years) of *PRKN*-PD onset.^17^ The motor symptoms were typical for *PRKN*-PD, presenting dyskinesia, motor fluctuations, and a good response to levodopa.^3^ Both of the patients did not present dysautonomia and cognitive decline, which is common for *PRKN*-PD.^3^ The older brother presented with impulse control disorder, which is one of the psychological symptoms that are not common, as only 2% of the *PRKN*-PD patients presents this.^17^ The older brother (III-1) performed STN-DBS operation with an improvement of motor symptoms. This finding was concordant with the literature on DBS suggesting that 76.1% of patients with *PRKN* variants displayed an excellent outcome with DBS.^39^

To date, over a hundred of *PRKN* causative variants (36 deletions, 22 duplications, 81 SNVs or indels, https://www.mdsgene.org/, Feb 2023) are reported and only one of these is an inversion including exon 2 to exon 5, which is different from the one identified in our study (c.8-224652_618+32307dupinvAAGATTTins). This inversion was identified from young onset PD patients in Poland.^32^ When searching Clinvar (https://www.ncbi.nlm.nih.gov/clinvar/?term=PRKN%5Bsym%5D), one inversion (c.618+7842_618+7934inv) in intron 5 was registered in *PRKN*. There was another case report describing this inversion in *PRKN*. A report from Israel describes the pathogenic homozygous inversion of exon 5 in *PRKN*.^40^ They report a consanguineous family with four out of 12 siblings developed young onset dystonia-parkinsonism. The symptoms were severe, with a median AAO of 11 years. Dystonia generalized gradually to the other limbs. A few years after the onset, they presented parkinsonism with motor fluctuation. Three out of four affected individuals had subthalamic deep brain stimulation. Two of the affected subjects had 18F-l-3,4-dihydroxyphenylalanine brain PET/CT studies and showed impaired striatal presynaptic dopaminergic function. Using short read WGS, the affected subjects were shown to harbor biallelic inversion including exon 5 followed by 49kb deletion, which causes skipping of exon 5 in the cDNA.

While the role of biallelic variants in *PRKN* is well established, the effects of heterozygous *PRKN* in PD are conflicting.^11–16^ Meta-analysis of *PRKN* heterozygous suggests an association between the heterozygous *PRKN* variant and PD. However, the potential confounding effect of an unrevealed second mutation is pointed out.^14^ Recently another study showed no association between heterozygous *PRKN* and PD by comparing short-read sequencing WGS.^16^ One potential explanation is that there could be other complex SVs that affect *PRKN* function or deep intronic variants which affect splicing that are missed in previous genotyping technologies. We speculate that some PD cases harboring heterozygous *PRKN* variants may have a complex inversion as a second variant, especially in the young-onset PD following an autosomal recessive pattern. As our cases had been classified as heterozygous *PRKN* carriers until the large inversion was identified, there are likely other young onset PD cases carrying unrevealed SVs, especially inversions or other complex SVs. Our previous report shows 2.5% of familial PD cases harbor *PRKN* heterozygous variants.^41^

Next, we surveyed large scale short-read WGS datasets to screen similar inversions in the general population including also PD cohorts. Overall, inversions of *PRKN* in both the UK Biobank and AMP-PD datasets were rare, and also we did not identify the exact inversion or a similar size inversion as the here reported *PRKN* family. However, all identified *PRKN* inversions are likely pathogenic and therefore of interest. It is interesting that the age of PD onset is relatively young (42 years) in the subject who harbored two inversions in AMP-PD dataset.Naturally the low frequency of *PRKN* inversions might be due inversions itself being quite rare in *PRKN* but it is also quite possible that short-read sequencing could not identify SVs. For example it is known that repeats or transposable elements can cause SVs and short read sequencing methods often struggle to map accurately in those regions.^42 43^

As with any study there are some limitations in the presented work. First, we did not have access to RNAseq data to assess the effect of the inversion on the transcript levels and isoforms. However, given the predicted structure of the inversion, it is likely that all *PRKN* isoforms including exon 12 will result in a non functioning transcript given the lack of a stop codon. We are planning to investigate this effect using differentiated neurons derived from iPSC. Second, the DNA of the father (II-2) was not available. However, as the non-affected mother (II-3) only harbors the large inversion, including *PRKN*, it is very likely that the father is carrier of the exon 3 deletion. Third, in our assessment of the frequency of potential damaging *PRKN* inversion in the general population we only included European ancestry individuals. Future studies should explore the frequency of *PRKN* inversion across populations.

In summary, here we report how long-read sequencing can identify complex *PRKN* SVs which are likely to be missed by MLPA and conventional short-read sequencing methods. We expect that several other early-onset PD cases with a *PRKN* phenotype have a second complex variant that long-read sequencing can resolve. This study emphasizes the usefulness of long-read sequencing in the research of familial PD cases.

## Data Availability

https://www.ukbiobank.ac.uk/

https://amp-pd.org/

## Acknowledgments

We would like to thank all of the subjects who donated their time and biological samples to be part of this study. This work was partly supported by the Intramural Research Program of the National Institute on Aging, the Intractable Disease Research Center of Juntendo University Graduate School of Medicine. We thank the Biowulf team, as this study used the high-performance computational capabilities of the Biowulf Linux cluster at the National Institutes of Health (http://hpc.nih.gov).

## Authors’ Roles

Study concept and design: K.Daida, M.Funayama, C.Blauwendraat, N.Hattori

Acquisition of data: K.Daida, M.Funayama, K.Billingsley, L.Malik, A.Miano-Burkhardt, H.Leonard, M.Makarious, H.Iwaki, J.Ding, J.Gibbs, M.Ishiguro, H.Yoshino, K.Ogaki, G.Oyama, K.Nishioka, R.Nonaka, W.Akamatsu

Writing the manuscript: K.Daida, M.Funayama, K.Billingsley, A.Miano-Burkhardt, C.Blauwendraat, N.Hattori

Editing the final version of the manuscript: K.Daida, M.Funayama, K.Billingsley, L.Malik, A.Miano-Burkhardt, H.Leonard, M.Makarious, H.Iwaki, J.Ding, J.Gibbs, M.Ishiguro, H.Yoshino, K.Ogaki, G.Oyama, K.Nishioka, R.Nonaka, W.Akamatsu, C.Blauwendraat, N.Hattori

