## Supplementary File for "Long-read sequencing resolves a complex structural variant in *PRKN* Parkinson’s disease"

**Supplementary material**

**Supplementary Methods**

**Supplementary Figure 1.** Gel image for electrophoresis of the PCR product around the breakpoints of inversion.

**Supplementary Figure 2.** IGV screenshots of the inversion in short-read whole genome sequencing.

**Supplementary Figure 3.** IGV screenshots of the deletion in short-read whole genome sequencing.

**Supplementary Figure 4.** Sample DNA QC results after size selection.

**Supplementary Figure 5.** IGV screen shots of inversions including PRKN exons identified in AMP-PD and UK biobank short-read whole genome sequencing.

**Supplementary Table 1.** Primer sequence of PCR around breakpoints

**Supplementary Table 2.** Summary for clinical symptoms of affected twins

**Supplementary Table 3.** Output data of long-read sequencing

**Supplementary Table 4.** Quality control of sample DNA

**Supplementary Table 5.** Inversions found in AMP-PD and UK Biobank

**Supplementary figure 1:** Gel image for electrophoresis of the PCR product around the breakpoints of inversion.

The figure at the top shows the gel with the PCR product using primers around 5’ breakpoint of the inversion. The figure at the middle shows the gel with the PCR product using primers around 3’ breakpoint of the inversion. The figure at the bottom shows the gel with the PCR product using 3' breakpoint primers of the reference.

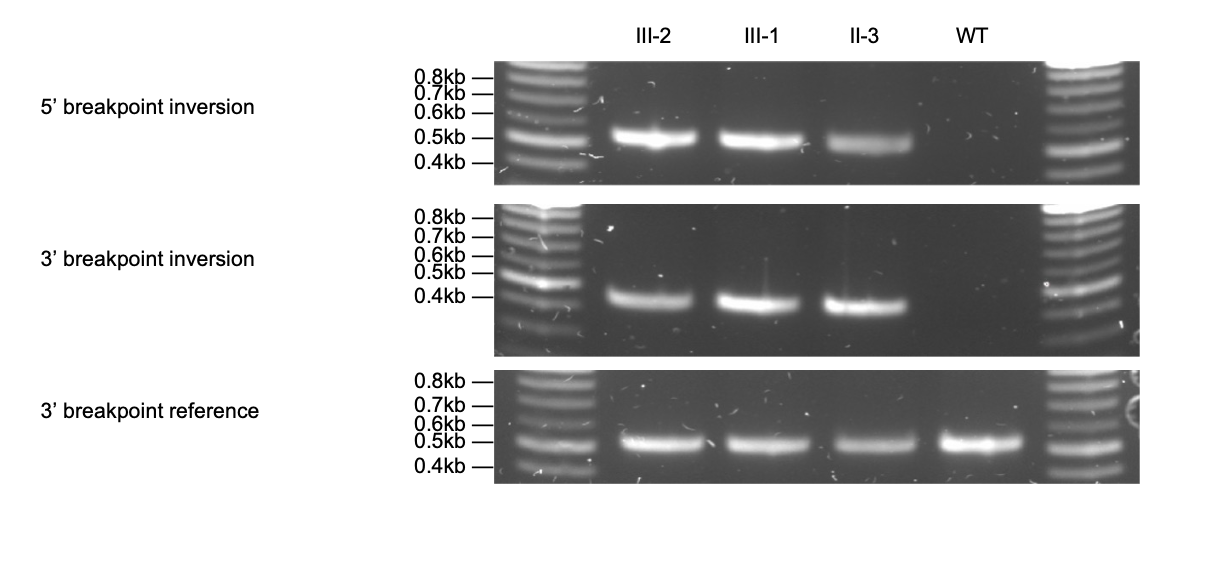

**Supplementary Figure 2:** IGV screenshots of the inversion in short-read whole genome sequencing

The upper figure shows the 5’ break point. The lower shows the 3’ break point.

WGS, whole genome sequencing; IGV, integrative genome viewer

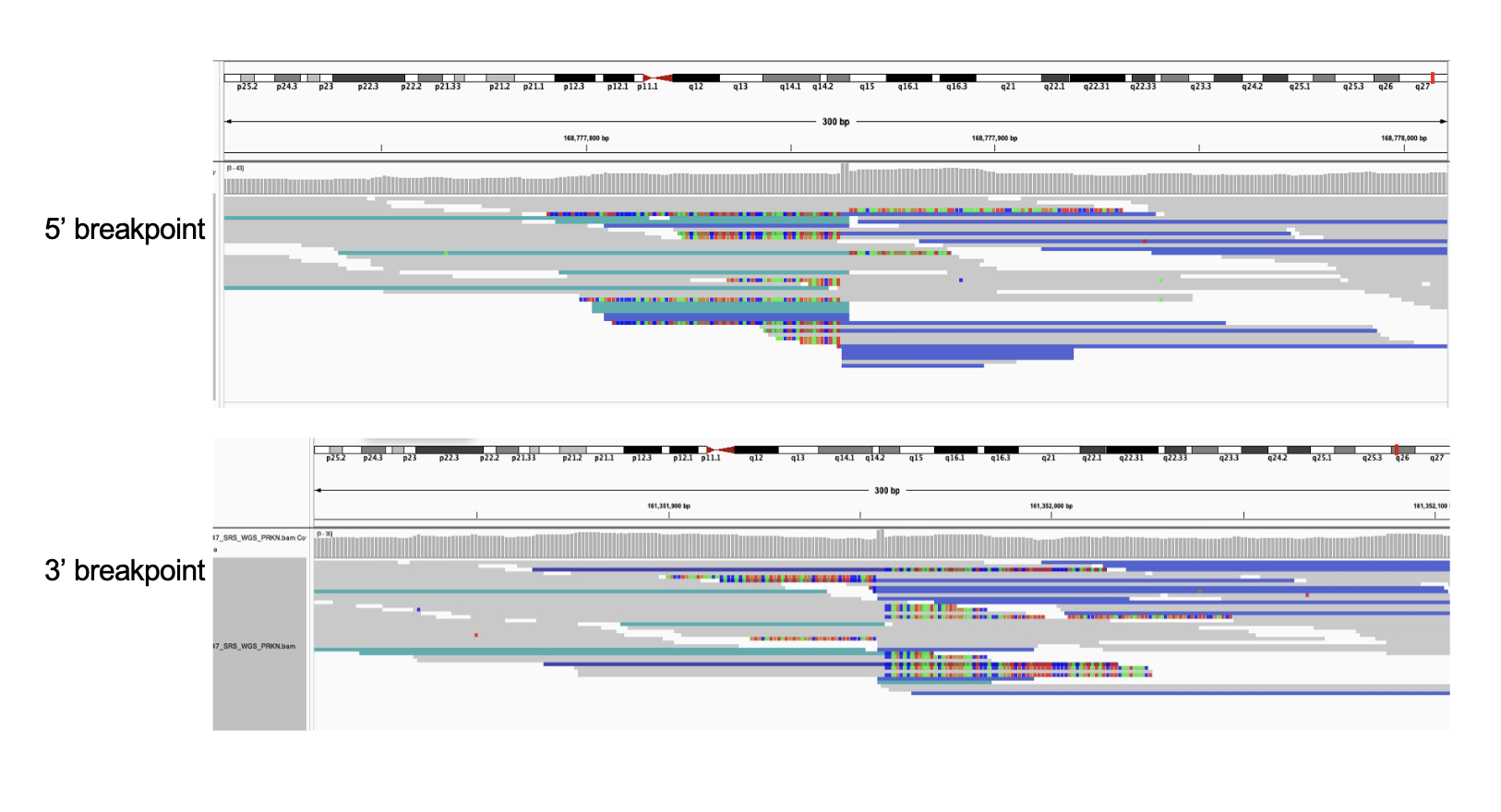

**Supplementary figure 3.** IGV screenshots of the deletion in short-read whole genome sequencing.

The upper figure shows the 5’ break point. The lower shows the 3’ break point.

WGS, whole genome sequencing

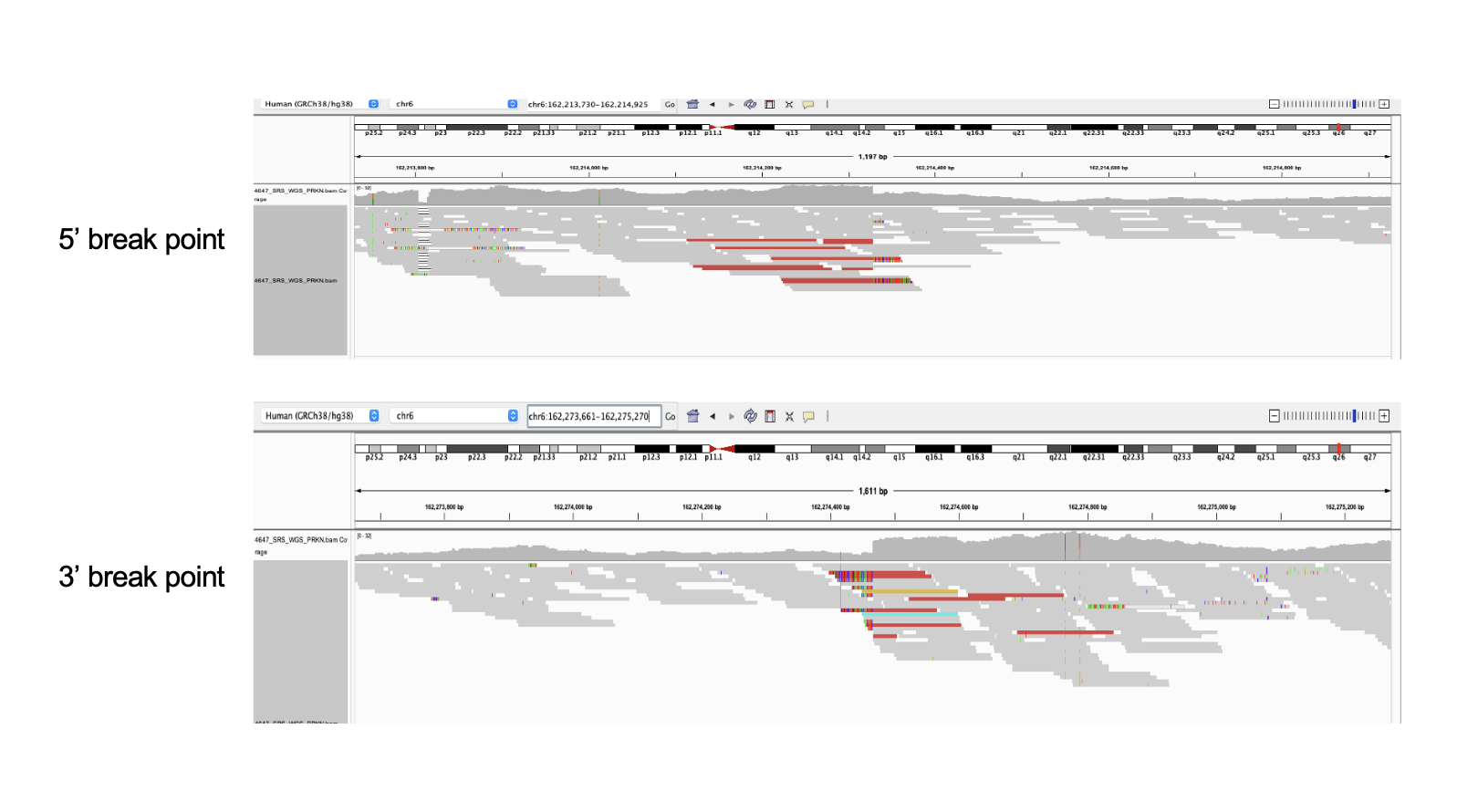

**Supplementary figure 4.** Sample DNA QC results after size selection.

DNA quality control results after size selection for long-read sequencing.

QC, quality control

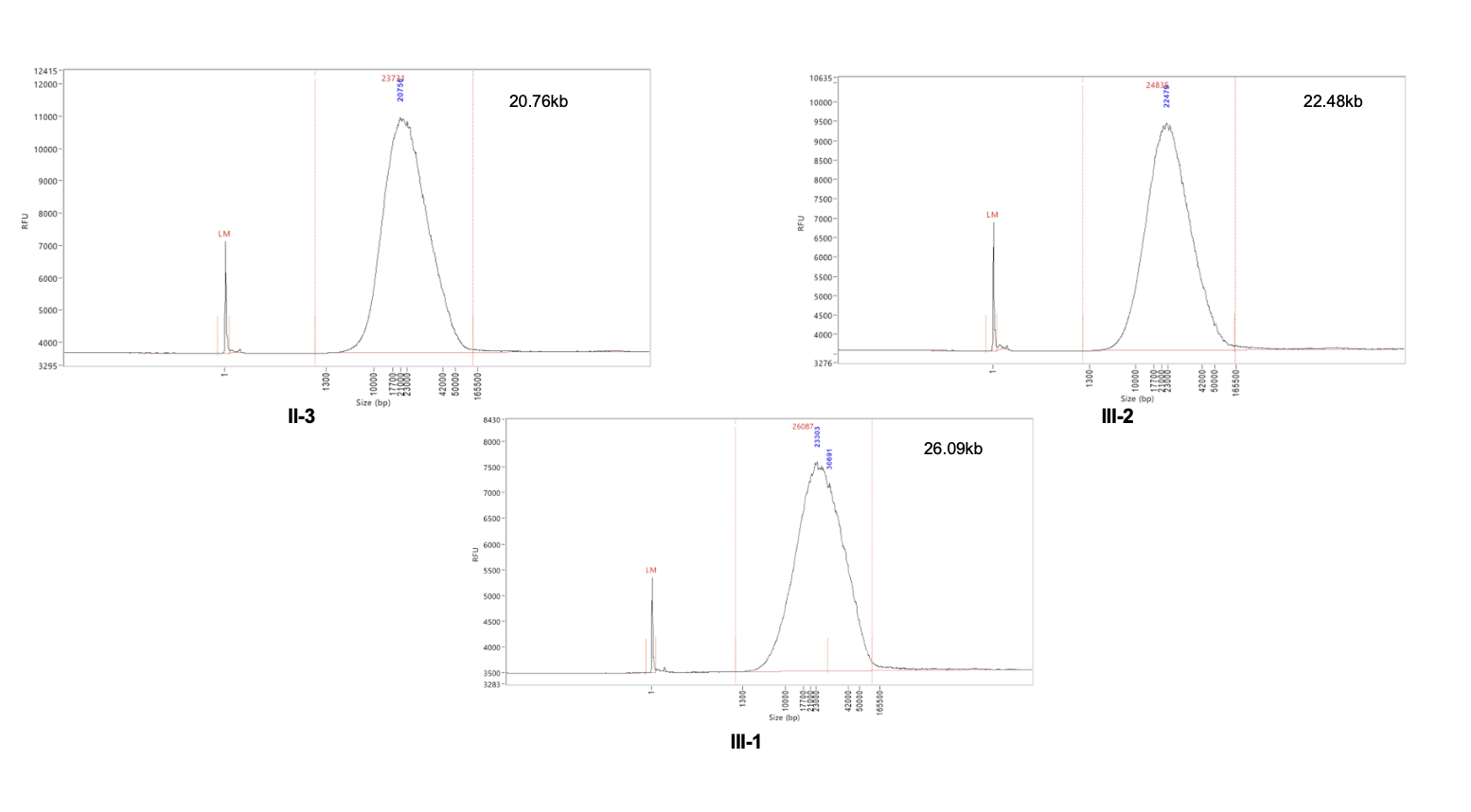

**Supplementary figure 5:** IGV screen shots of inversions including PRKN exons identified in AMP-PD and UK biobank short-read whole genome sequencing.

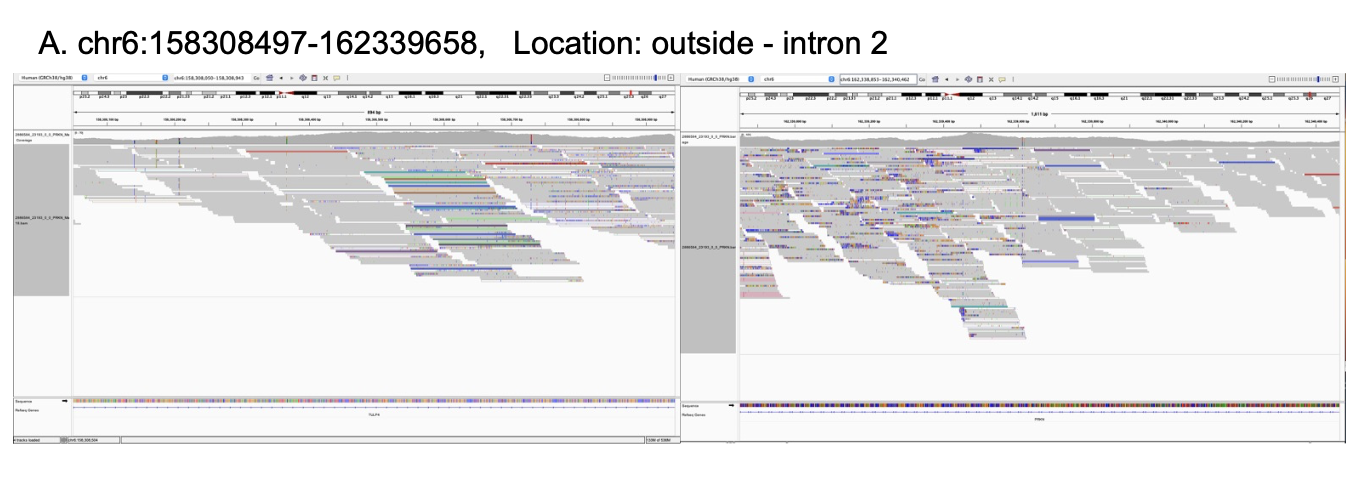

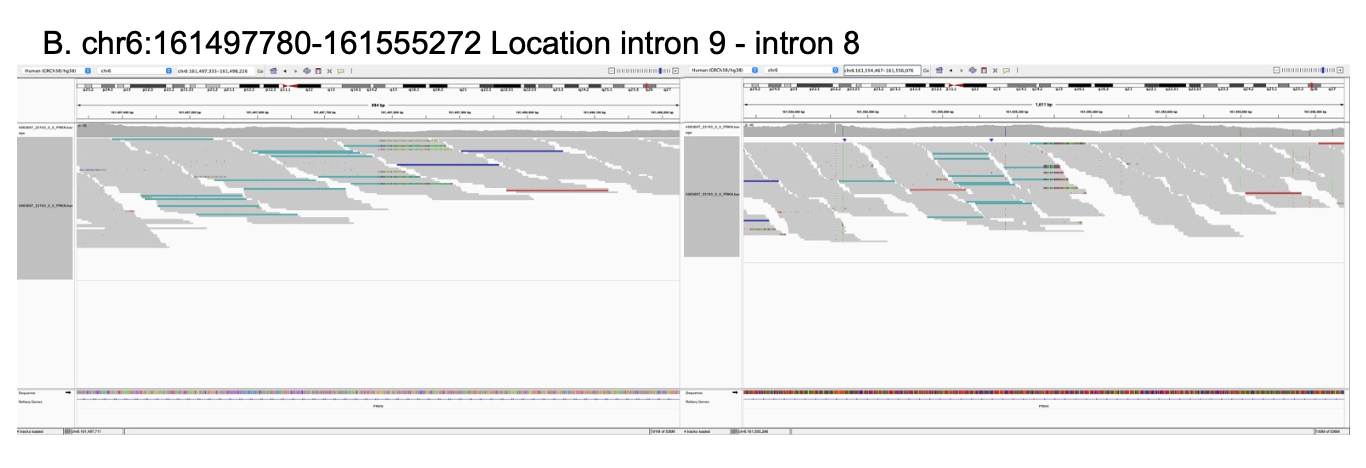

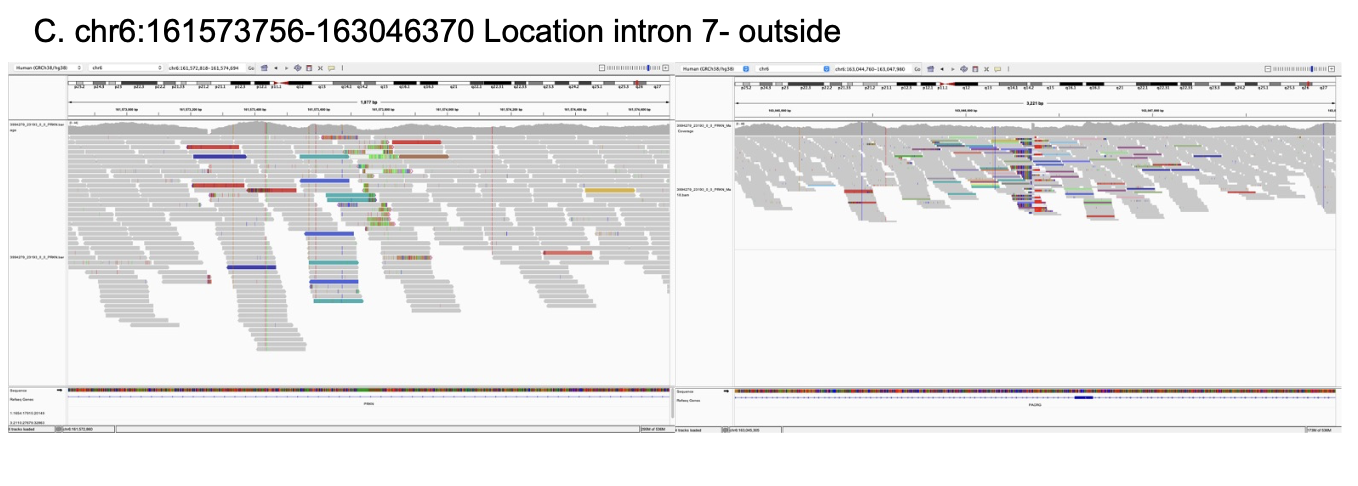

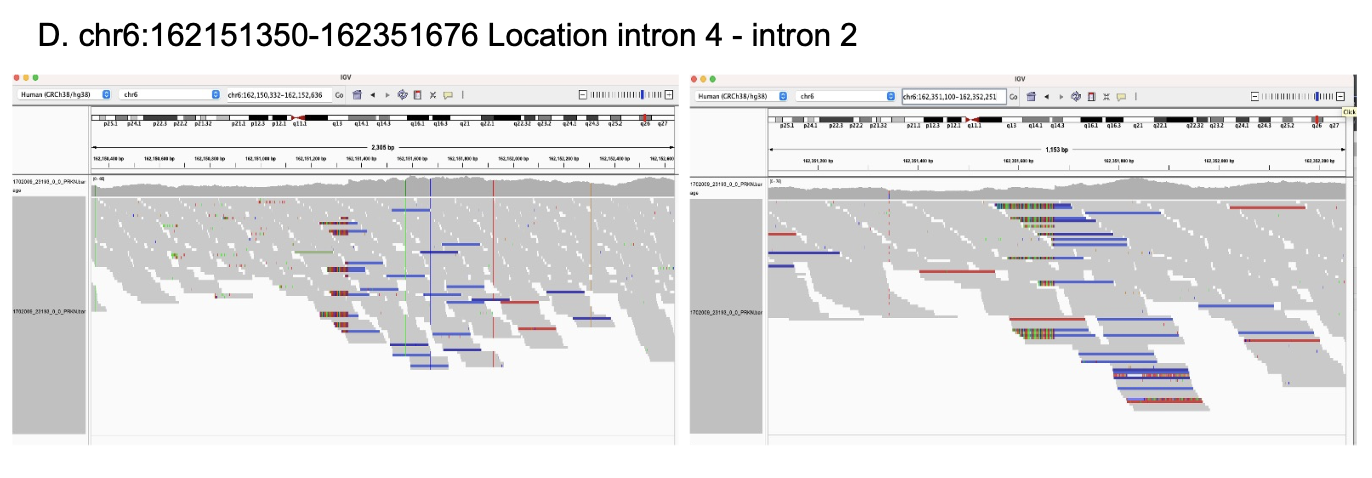

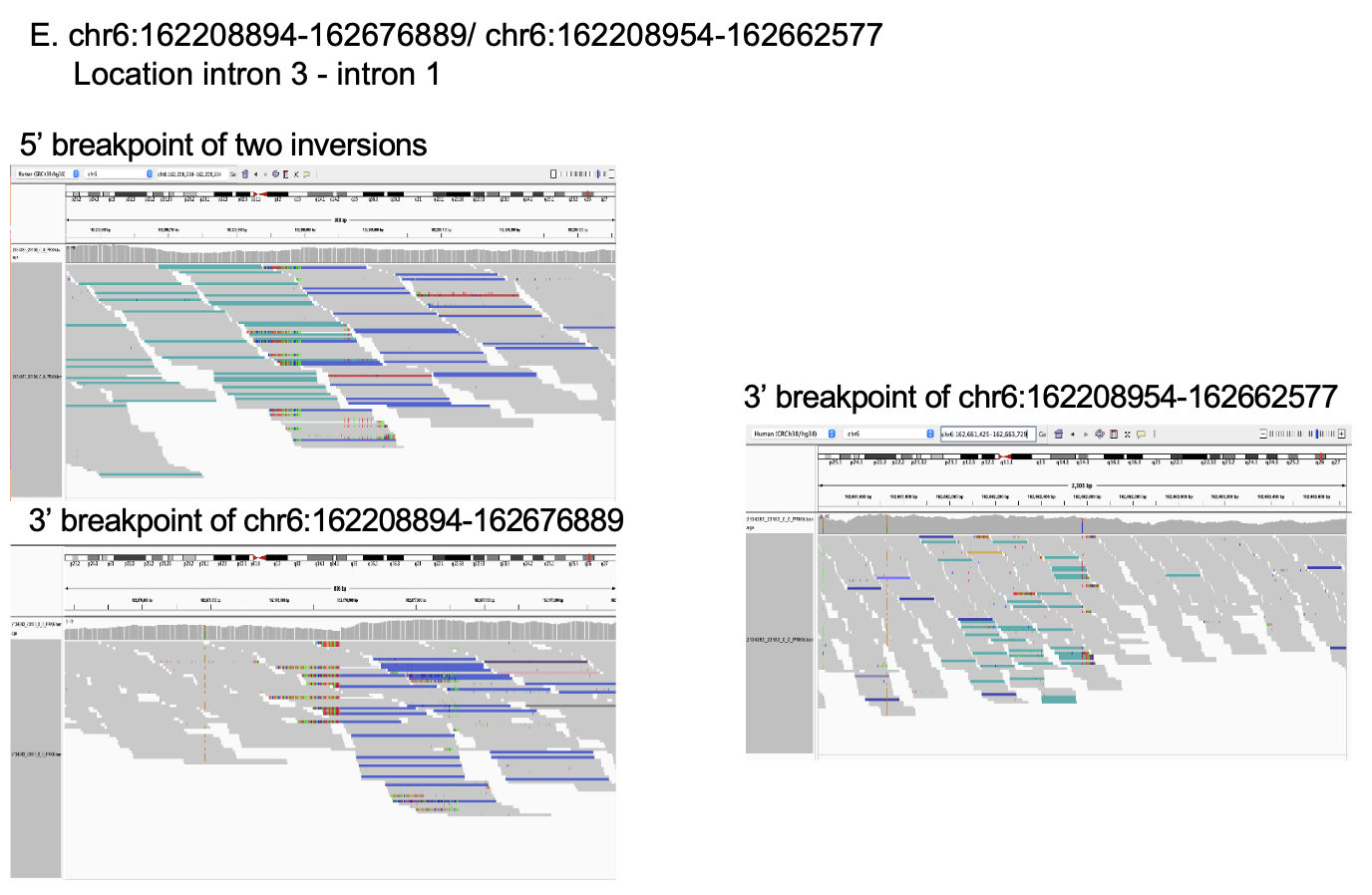

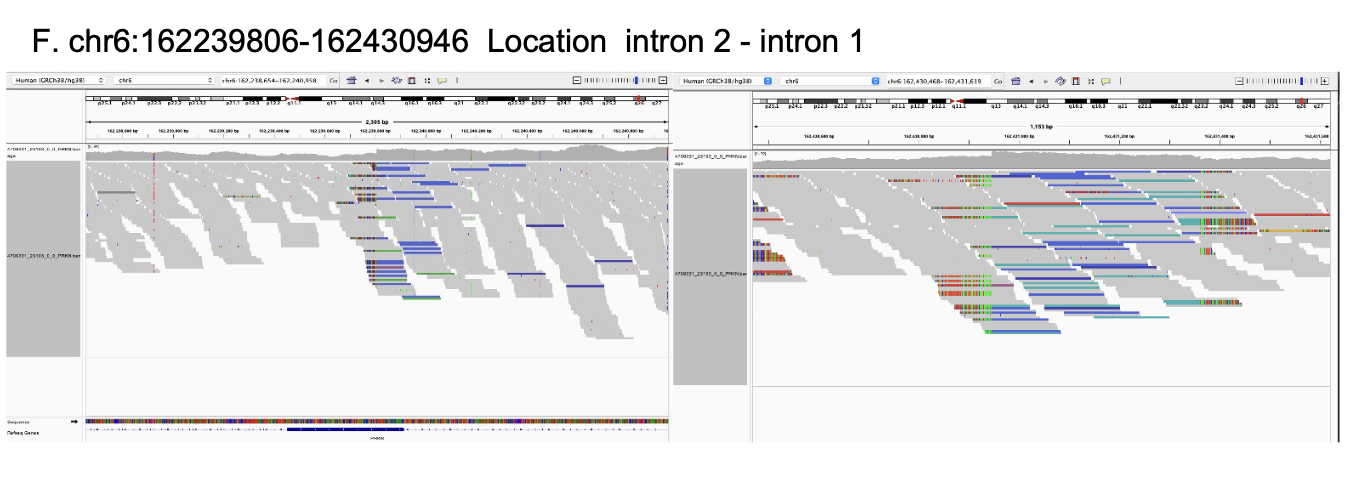

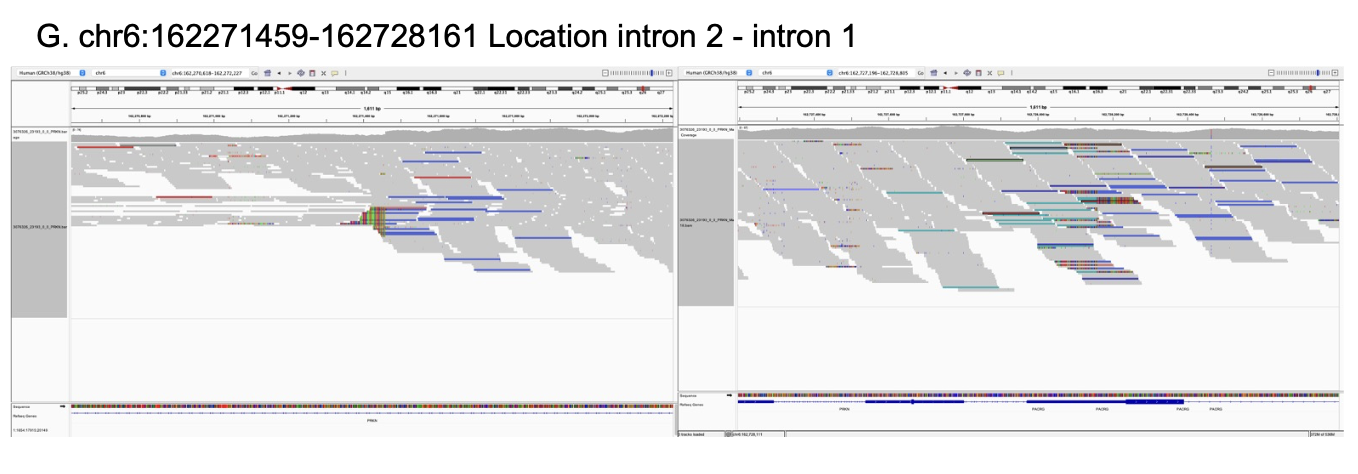

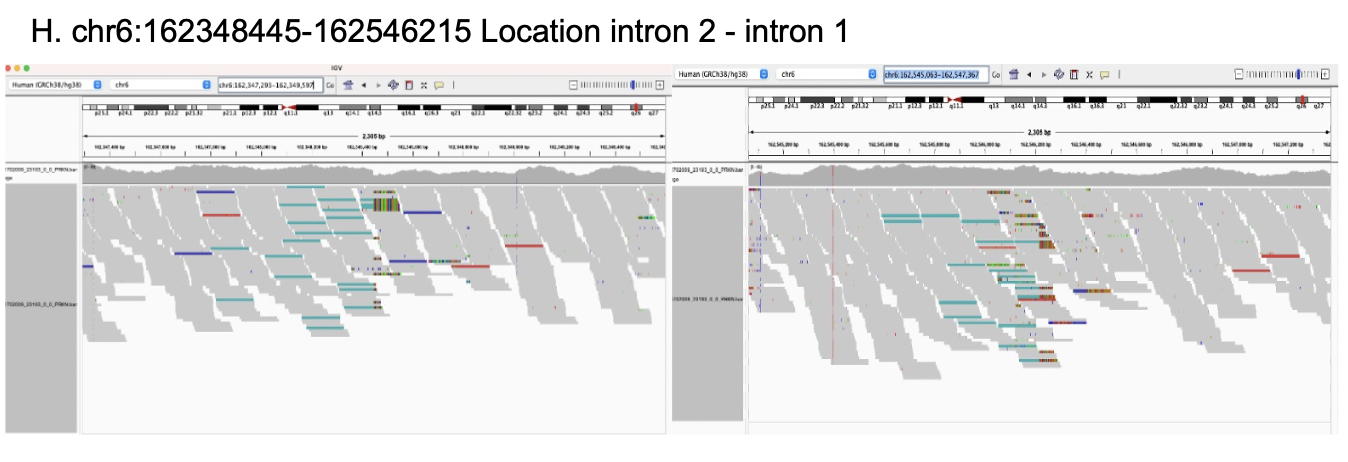

**Supplementary Table 1: Primer sequence of PCR around breakpoints**

| Target region | Forward | Reverse |
| --- | --- | --- |
| breakpoint 1 with inversion | CACCCAGCCAGCATATTATTTT | AGAGCCCGGGATAATGTGTC |
| breakpoint 2 with inversion | GAGACAGCACCGAACCAATT | AATGCTTGCTGTCCACATCC |
| breakpoint 2 of reference (hg38) | ACTGGAGAGCCCGGGATAAT | CCGACCTCCTAGCTGTCTCT |

**Supplementary Table 2: Summary for clinical symptoms of affected twins**

AAO, age at onset; AAE, age at examination; F, female; M, male; MMSE, Mini-Mental State Examination; REM, rapid eye movement; MRI, magnetic resonance imaging; DAT-SPECT, dopamine transporter single-photon emission computed tomography with 123I-ioflupane; MIBG, 123iodine- metaiodobenzylguanidine myocardial scintigraphy; UPDRS, Unified Parkinson’s Disease Rating Scale; N/A, not available.

This table is accessible upon request to the corresponding author.

**Supplementary Table 3:** Output data of long-read sequencing

Data output from Oxford nanopore sequencer PromtehION.

Gb, giga base

| **SAMPLE** | **Overall output (Gb)** | **Read N50 (kb)** | **Estimated Coverage** |
| --- | --- | --- | --- |
| II-3 | 118.77 | 18.99 | 36.97 |
| III-1 | 117.61 | 19.3 | 37.00 |
| III-2 | 133.98 | 19.71 | 38.98 |

**Supplementary Table 4: Quality control of sample DNA**

|  | Qubit | | Nanodrop | | | Femto Pulse |
| --- | --- | --- | --- | --- | --- | --- |
| Sample | ng/ul | ug | 260/280 | 260/230 | ng/ul | Femto Peak (kb) |
| II-3 | 35.2 | 3.41 | 1.96 | 1.82 | 41.46 | 20.76 |
| III-1 | 42.8 | 4.15 | 1.8 | 1.91 | 54.59 | 26.09 |
| III-2 | 36.2 | 3.48 | 1.91 | 1.62 | 43.3 | 22.48 |

**Supplementary Table 5: Inversions found in AMP-PD and UK Biobank**

Details of the inversions including *PRKN* exons identified in AMP-PD and UK Biobank short-read whole genome sequencing datasets. Carriers in AMP-PD and Carriers in UKB indicates the number of subjects identified in each dataset and numbers in parentheses indicate the number of those with Parkinson’s disease. Location column shows the location of the inversion in *PRKN*.

PD, Parkinson’s disease; IGV, integrative genome viewer

| **chr** | **start** | **end** | **type** | **Size (bp)** | **Carriers in AMP-PD (carriers with PD)** | **Carriers in UKB (carriers with PD)** | **IGV false postive** | **Location** |
| --- | --- | --- | --- | --- | --- | --- | --- | --- |
| chr6 | 158308497 | 162339658 | INV | 4031161 | 0 (0) | 1 (0) | + | outside-intron2 |
| chr6 | 161497780 | 161555272 | INV | 57492 | 0 (0) | 1 (0) | - | intron 9 - intron 8 |
| chr6 | 161573756 | 163046370 | INV | 1472614 | 0 (0) | 1 (0) | - | intron 7- outside |
| chr6 | 162151350 | 162351676 | INV | 200326 | 0 (0) | 1 (0) | - | intron 4 - intron 2 |
| chr6 | 162208894 | 162676889 | INV | 467995 | 1 (1) | 1 (0) | - | intron 3 - intron 1 |
| chr6 | 162208954 | 162662577 | INV | 453623 | 1 (1) | 1 (0) | - | intron 3 - intron 1 |
| chr6 | 162239806 | 162430946 | INV | 191140 | 0 (0) | 6 (0) | - | intron 2 - intron 1 |
| chr6 | 162271459 | 162728161 | INV | 456702 | 0 (0) | 1 (0) | - | intron 2 - intron 1 |
| chr6 | 162348445 | 162546215 | INV | 197770 | 0 (0) | 1 (0) | - | intron 2 - intron 1 |
